# Characterizing altruistic motivation in potential volunteers for SARS-CoV-2 challenge trials

**DOI:** 10.1101/2021.03.14.21253548

**Authors:** Sophie M Rose, Virginia L Schmit, Thomas C Darton, Nir Eyal, Monica Magalhaes, Josh Morrison, Matthew Peeler, Seema K Shah, Abigail A Marsh

## Abstract

In human challenge trials, volunteers are deliberately infected with a pathogen to accelerate vaccine development and answer key scientific questions. In the U.S., preparations for challenge trials with the novel coronavirus are complete, and in the U.K., challenge trials have recently begun. However, ethical concerns have been raised about the potential for invalid consent or exploitation. These concerns largely reflect worries that challenge trial volunteers may be unusually risk-seeking or too economically vulnerable to refuse the payments these trials provide, rather than being motivated primarily by altruistic goals. We conducted the first large-scale survey of intended human challenge trial volunteers and found that SARS-CoV-2 challenge trial volunteers exhibit high levels of altruistic motivations without any special indication of poor risk perception or economic vulnerability. Findings indicate that challenge trials with the novel coronavirus can attract volunteers with background conditions, attitudes, and motivations that should allay key ethical concerns.

---

The ongoing COVID-19 pandemic presents extraordinary threats to public health and human welfare. Economic and social recoveries will require ongoing development and testing of prevention strategies, including vaccines that are easier to provide, store, and deliver; various dosing regimens, and updating of vaccines to keep pace with emerging mutations^1^. Human challenge trials, in which volunteers are deliberately infected (or “challenged”) with the pathogen to test vaccine candidates, are among the most efficient and scientifically powerful approaches to testing vaccines and learning about early disease processes^2^. Well-designed human challenge trials can speed the development of improved vaccines by selecting the most promising candidates to prioritize for further testing^3,4,5,6,7,8^.

The potential for benefits from challenge trials are largely societal. By contrast, the risks and burdens of challenge trials—including infection-related risks, prolonged period of biocontainment and possible trial vaccine or treatment side effects—fall largely on volunteers^9^. These risks and burdens (which are heightened by uncertainty about COVID-19 disease outcomes) coupled with the absence of obvious direct benefits for volunteers have led some bioethicists to suggest that challenge trials using the novel coronavirus may be unethical^10,11,12^. Some commentators worry that challenge trials might attract volunteers who are vulnerable to undue inducement or problems understanding relevant risks, which might invalidate volunteers’ consent or result in their exploitation^13,14^.

Addressing ethical concerns is made all the more pressing now that a COVID-19 human challenge trial has recently begun in the United Kingdom^15^. However, there are limited data on volunteer motivations and understanding for participation in human challenge trials, and none on volunteers willing to participate in challenge trials with the novel coronavirus^16,17,18^.

Direct benefits to participants are not required for human subjects research to be considered ethical^19^. Instead, the totality of the benefits—including benefits to others—should be sufficient to justify the risks. Trials also should be designed to expose participants to as few risks as possible, and participants must be able to provide valid informed consent^20^. This requires providing volunteers with the opportunity to evaluate the risks, benefits, and alternatives to any intervention to ensure that it reflects their goals, preferences, and values^21^.

Given the altruistic nature of challenge trial participation—with volunteers required to take on personal risks and costs to achieve societal benefits—it would be ideal from an ethical perspective if volunteers demonstrated highly altruistic goals, values and preferences. To date, few studies have examined why healthy volunteers consent to research with net risks and burdens to themselves, or whether their goals and values are compatible with ethical participation^16,17,22,23,24,25^. To assess whether a group of individuals who proactively declared their intent to volunteer to participate in a COVID-19 challenge trial meets these conditions, we conducted the first large-scale evaluation of characteristics of potential challenge trial volunteers. Volunteers were recruited through the non-profit advocacy organization 1Day Sooner (https://www.1daysooner.org/). 1Day Sooner was created in April 2020 to accelerate the deployment of effective vaccines by supporting preparation efforts for COVID-19 challenge trials and to advocate on behalf of COVID-19 human challenge trial volunteers. It curates the only centralized international database of volunteers who have indicated their willingness to partake in COVID-19 challenge trials.

We hypothesized that COVID-19 challenge trial volunteerism reflects heightened altruistic values and preferences. In light of concerns that challenge trials may attract participants who are unusually insensitive to risk or who are in dire economic need^10,11,17^, we also tested the alternate hypotheses that challenge trials attract participants who engage in elevated risk behaviors (including specifically health and safety related risk behaviors) or those who are economically or otherwise vulnerable to exploitation. Either of these issues could raise concerns about the ethical permissibility of trials, although trialists could still try to select for those intended volunteers who have accurate risk perceptions and no socioeconomic motivations.

To test these hypotheses, we conducted a pre-registered (https://osf.io/fqyrb) study in which we measured altruistic motivation, values, and behavior; risk preferences and behaviors, and sociodemographic variables in 1,911 potential COVID-19 challenge trial volunteers. We compared volunteers to 999 controls recruited to reflect approximate 2019 US Census demographics, whose characteristics are described in Table 1.

**Table 1:** Participant demographic characteristics.

|  | **Volunteer Group**<br>**(n=1911)**<br>*N.o. people (%)* | **Control**<br>**(n=999)**<br>*N.o. people (%)* |
| --- | --- | --- |
| **Age** |  |  |
| 18-25 | 252 (13.3) | 91 (9.4) |
| 26-35 | 580 (30.7) | 139 (14.4) |
| 36-45 | 419 (22.2) | 195 (20.1) |
| 46-55 | 297 (15.7) | 189 (19.5) |
| 56-65 | 229 (12.1) | 139 (14.4) |
| 66-75 | 99 (5.2) | 180 (18.6) |
| 76+ | 12 (0.6) | 35 (3.5) |
| Non-responses | 23 | 31 |
| **Gender** |  |  |
| Male | 1151 (60.4) | 436 (43.9) |
| Female | 673 (35.3) | 522 (52.5) |
| Self-identify/Prefer not to say | 82 (4.3) | 36 (3.6) |
| Non-response | 5 | 5 |
| **Marital Status** |  |  |
| Single (never married) | 976 (51.1) | 289 (29.1) |
| Married/Domestic partnership | 664 (34.7) | 523 (52.6) |
| Divorced | 200 (10.5) | 121 (12.2) |
| Widowed | 27 (1.4) | 48 (4.8) |
| Separated | 44 (2.3) | 13 (1.3) |
| Non-responses | 0 | 5 |
| **Race/Ethnicity** |  |  |
| Selected African (Yes/No (%Yes)) | 28/1883 (1.5) | 95/904 (9.5) |
| Selected Hispanic (Yes/No (%Yes)) | 133/1778 (7.0) | 59/940 (5.9) |
| Selected Caucasian (Yes/No (%Yes)) | 1595/316 (83.5) | 706/293 (70.7) |
| Selected Asian (Yes/No (% Yes)) | 162/1749 (8.5) | 110/889 (12.4) |
| Selected Native American (Yes/No (% Yes)) | 32/1879 (1.7) | 19/980 (1.9) |
| **Employment (Top 5 categories listed)** |  |  |
| Employed | 1009 (52.8) | 394 (39.4) |
| Self-employed/Freelance | 197 (10.3) | 52 (5.2) |
| Retired | 137 (7.2) | 240 (24.0) |
| Studying | 121 (6.3) | 36 (3.6) |
| Unemployed/Looking for work | 115 (6.0) | 80 (8.0) |
| **Employment Status** |  |  |
| Employed full-time | 941 (49.3) | 323 (32.6) |
| Unemployed | 728 (38.2) | 593 (59.9) |
| Employed part-time | 239 (12.5) | 74 (7.5) |
| Non-responses | 3 | 9 |
| **Income (in USD)** |  |  |
| Less than \$25K | 211 (11.0) | 217 (21.8) |
| \$25K-\$50K | 326 (17.1) | 248 (24.9) |
| \$50K-\$100K | 512 (26.8) | 203 (20.4) |
| \$100K-\$200K | 458 (24.0) | 240 (24.1) |
| Greater than \$200K | 260 (13.6) | 67 (6.7) |
| Prefer not to say | 143 (7.5) | 21 (2.1) |
| Non-responses | 1 | 3 |
| **Have health insurance** |  |  |
| Yes | 1683 (88.1) | 877 (88.1) |
| No | 180 (9.4) | 95 (9.5) |
| Not sure | 29 (1.5) | 10 (1.0) |
| Prefer not to say | 18 (0.9) | 14 (1.4) |
| Non-responses | 1 | 3 |
| **Have Children (Yes/No (% Yes))** | 549/1360 (28.8) | 494/499 (49.7) |
| Non-responses | 2 | 6 |

| **Number in Household (other than self)** |  |  |
| --- | --- | --- |
| 0 | 611 (32.2) | 267 (27.0) |
| 1 | 656 (34.5) | 353 (35.7) |
| 2 | 260 (13.7) | 152 (15.4) |
| 3 | 228 (12.0) | 116 (11.7) |
| 4 | 89 (4.7) | 73 (7.4) |
| 5 | 34 (1.8) | 21 (2.1) |
| 6 | 11 (0.6) | 5 (0.5) |
| 7 | 2 (0.1) | 2 (0.2) |
| 8+ | 9 (0.5) | 1 (0.1) |
| Non-responses | 11 | 9 |
| **Education Level** |  |  |
| Less than High School | 4 (0.2) | 22 (2.2) |
| High School Graduate/GED | 84 (4.4) | 163 (16.3) |
| Some college, no degree | 218 (11.4) | 149 (14.9) |
| Trade/Technical training | 65 (3.4) | 42 (4.2) |
| Associate degree | 59 (3.1) | 102 (10.2) |
| Bachelor's degree | 684 (35.8) | 280 (28.1) |
| Master's degree | 498 (26.1) | 172 (17.3) |
| Professional degree | 119 (6.2) | 44 (4.4) |
| Doctoral degree | 180 (9.4) | 23 (2.3) |
| Non-responses | 0 | 2 |

## Results

### Socio-demographic variables

Most volunteers (66.2%) were between 18 and 45 years of age, identified as non-Hispanic white (78.5%), and had a bachelor’s degree or higher (77.4%). A majority reported residing in the United States (81.5%), followed by Canada (7.6%), the United Kingdom (2.3%) and Germany (1.0%). Most volunteers had either private health insurance or access to healthcare through publicly-funded health systems (88.1%). Approximately one in three volunteers (32.0%) lived alone, and a similar proportion (34.4%) lived with only one other person. 28.7% of volunteers had at least one child. Half of volunteers were employed full-time (50.8%) and most (71.9%) reported an annual household income greater than US $50,000. Of the total, 213 reported an annual household income less than US $25,000 (see Table 1); 23% of these (49/213) were students.

Comparing the two groups, more volunteers were male (60.4%, N=1151) relative to the general population and our controls (43.9%, N=439). Of volunteers, 35.3% self-identified as female (N=680); 3.2% self-identified as non-binary or transgender (N=61), and 1.1% did not specify their gender (N=21). Volunteers were generally younger (44.0% under age 35 versus 23.8% of controls) and more educated (77.5% reported earning a Bachelor’s degree or higher, versus 52.1% of controls). Volunteers were also wealthier; assuming equal distribution within income categories, 61.9% of volunteers were above the U.S. median income ($68,703 annually), compared to 45.7% of the control group. Of volunteers, 11.8% fell below the U.S. poverty line ($26,172 annually for a family of four), compared to 23.0% of controls^26^. Volunteers and controls reported equal levels of health insurance (88.1% of both groups).

### Altruistic values and preferences

Following our pre-registered analysis plan, we conducted an exploratory factor analysis on responses to the 10 motivations for volunteering (see Supplementary Information), which returned a three factor solution, with one factor comprising the two altruistic motivations. The percentages of participants who selected each of the motivations were calculated (Table 2). The two altruistic motivations were the only options selected by majorities of respondents; both were selected by over three-quarters of volunteers (“I wanted to help others and potentially save lives” (95.9%) and “I wanted to contribute to the progress of medicine” (79.2%)). The third most highly ranked choice (“I feel helpless and this is a way to do something positive” (46.6%)) was selected by a minority of volunteers, as were the remaining options.

**Table 2:** Volunteer group motivations for participating in human challenge studies.

| Motivation | Number (%) rating motivation in the top three reasons for volunteering<sup>1</sup> |
| --- | --- |
| I wanted to help others and potentially save lives | 1832 (95.9) |
| I wanted to contribute to the progress of medicine | 1513 (79.2) |
| I feel helpless and this is a way to do something positive | 890 (46.6) |
| Another factor not mentioned | 380 (19.9) |
| I wanted to be part of a clinical trial | 348 (18.2) |
| I am likely to be infected by COVID-19 anyway | 282 (14.8) |
| I was curious about COVID-19 | 170 (8.9) |
| I wanted to be guaranteed access to critical care should I be infected with COVID-19 | 156 (8.2) |
| I wanted to find out more about my own health | 83 (4.3) |
| I wanted to receive the financial reimbursement for participating | 79 (4.1) |
| <sup>1</sup> Since volunteers were asked to rate whether the choices above were in their top three reasons, percentages total 300% instead of 100% (with exceptions due to rounding). |  |

We next conducted chi-square tests to compare challenge trial volunteers’ and controls’ prior engagement in altruistic behavior and found that volunteers were more likely than controls to have participated in all but one of these behaviors (Figure 1). More volunteers reported having previously donated blood (V: 75.5%, C: 62.5%, χ^2^(1)=54.020, *p*<0.001), having donated significant amounts of money to charity (V: 75.3%, C: 50.3%, χ^2^(1)=175.374, *p*<0.001), registering as a bone marrow donor (V: 35.5%, C: 14.7%, χ^2^(1)=124.284, *p*<0.001) or being a registered deceased organ donor (V: 85.8%, C: 47.4%, χ^2^(1)=460.221, *p*<0.001). More controls reported being living kidney or liver donors (V: 1.2%, C: 9.6%, χ^2^(1)=116.813, *p*<0.001), but positive response rates for controls were implausibly high given the overall prevalence of living organ donation (per capita prevalence < 1 in 100,000), suggesting results for this question may not be reliable.

**Figure 1.**
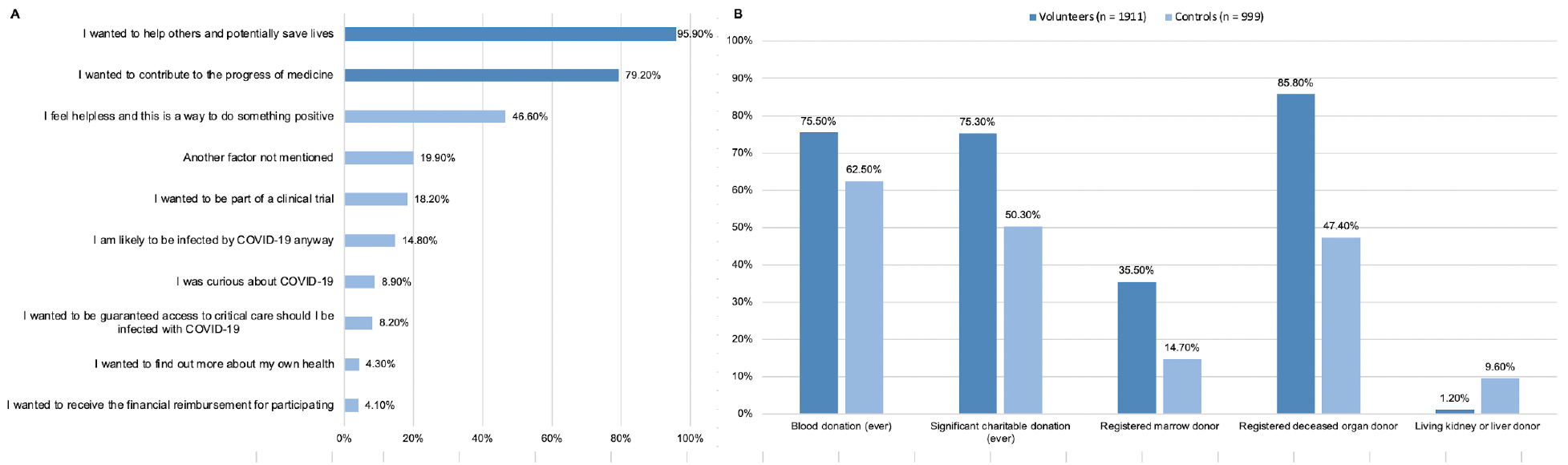
Volunteer group motivations for participating in human challenge studies and comparison of engagement in altruistic behaviors by volunteer vs. control groups.

We next compared volunteer and control groups along each HEXACO dimension using an ANCOVA model, controlling for age, income, education level, gender and country of residence. Effect sizes were calculated using eta-squared (η^2^) values^27^, with effect sizes <0.01 considered trivial, effect sizes 0.01-0.06 small, effect sizes 0.06-0.14 medium, and effect sizes >0.14 large. Average scores for volunteers were significantly higher than controls on all but one of the HEXACO dimensions, with the largest effect sizes obtained for Honesty-Humility (V: 4.25, C: 3.67, p<0.001, η^2^=0.128) and Openness to Experience (V: 3.96, C: 3.44, p <0.001, η^2^=0.119). In contrast, volunteers scored lower on Emotionality, but this effect size was small (V: 2.55, C: 2.84, p=0.03, η^2^ < 0.03) (Table 3).

**Table 3:** Comparisons of HEXACO dimension scores by volunteer vs. control group membership.

| | Volunteer Group<br>(n=1911) | | Control<br>(n=999) | | p-value | $\eta^2$ |
| --- | --- | --- | --- | --- | --- | --- |
|  | Mean (SE) | 95% CI | Mean (SE) | 95% CI |  |  |
| **Honesty-Humility (H)** | 4.249 (0.015) | 4.219, 4.278 | 3.672 (0.022) | 3.629, 3.715 | <0.001 | 0.128 |
| **Emotionality (E)** | 2.554 (0.016) | 2.523, 2.585 | 2.840 (0.023) | 2.795, 2.885 | <0.001 | 0.032 |
| **EXtraversion (X)** | 3.915 (0.016) | 3.883, 3.947 | 3.483 (0.024) | 3.436, 3.530 | <0.001 | 0.065 |
| **Agreeableness (A)** | 3.071 (0.014) | 3.044, 3.099 | 2.918 (0.021) | 2.878, 2.958 | <0.001 | 0.012 |
| **Conscientiousness (C)** | 3.727 (0.016) | 3.697, 3.758 | 3.498 (0.023) | 3.453, 3.543 | <0.001 | 0.021 |
| **Openness to** | 3.964 (0.014) | 3.936, 3.991 | 3.442 (0.021) | 3.402, 3.483 | <0.001 | 0.119 |
| **Experience (O)** |  |  |  |  |  |  |
| Age was considered a covariate and the other categorical variables were treated as fixed effects. Marginal means by group are the means for the two groups controlled for all covariates (assuming the mean age of the sample of 43.58). P-values were calculated using F-tests. Effect sizes were calculated using eta-squared ($\eta^2$), with cutpoints for small, medium, and large effects defined as 0.01, 0.06, and 0.14 respectively. |  |  |  |  |  |  |

We used logistic regression analyses to predict the likelihood of a participant being in the challenge study volunteer group based solely on HEXACO outcomes. Results indicated that volunteer status was most strongly predicted by openness to experience (OR: 4.60, 95% CI: 3.91, 5.41, d=0.841) when controlling for the five other HEXACO dimensions. Honesty-Humility was the next most strongly associated with volunteer group membership (OR: 2.36, 95% CI: 2.05, 2.72, d=0.473). Other dimensions were less strongly associated (effect sizes 0.101-0.238)^27^. This model had a Cox & Snell R^2^ = 0.246 and an Akaike Information Criterion (AIC) value of 2751.09^28^.

We then added demographic covariates to the above model, including age, gender, education level, income, and country of residence to control for the potential influence of these differences between volunteers and controls. Results indicate that, after controlling for these variables, volunteer status was most strongly associated with Openness to Experience (OR: 4.32, 95% CI: 3.53, 5.29, Cohen’s d=0.806) and Honesty-Humility (OR: 4.28, 95% CI: 3.52, 5.20, Cohen’s d=0.801) (Table 4 and Supplemental Table 1). Agreeableness was not associated with group membership (OR: 0.99, 95% CI: 0.81, 1.21). Effect sizes for other HEXACO dimensions ranged from 0.175 to 0.238. In addition, education level and income were both found to be significantly associated with volunteer group membership with a large effect size (Table 5). For example, study participants with an education level equivalent to a Bachelor’s degree had 144-fold odds of being a member of the volunteer group compared to those with less than high school equivalent education (OR: 144.14, p<0.001, d=2.74). Participants with an annual income of greater than $200,000 had 4.19-fold odds of being a member of the volunteer group compared to those earning less than $25,000 annually. This model had a total Cox & Snell R^2^ = 0.432 (with HEXACO dimension covariates accounting for 16.4% of R^2^) and an AIC value of 2070.92, indicating that the addition of demographic covariates improved the fit of the model overall.

**Table 4:** Odds of challenge volunteer membership by HEXACO dimension using logistic regression model, adjusted for gender, age, education, country of residence and income.

| HEXACO Dimension | Wald statistic | p-value | Odds Ratio | 95% CI for OR | Cohen's d |
| --- | --- | --- | --- | --- | --- |
| **Honesty/Humility** | 214.268 | < 0.001 | 4.278 | 3.521, 5.197 | 0.801 |
| **Emotionality** | 15.675 | < 0.001 | 0.684 | 0.567, 0.826 | 0.209 |
| **eXtraversion** | 22.814 | < 0.001 | 1.539 | 1.289, 1.836 | 0.238 |
| **Agreeableness** | 0.012 | 0.912 | 0.989 | 0.811, 1.206 |  |
| **Conscientiousness** | 10.309 | 0.001 | 0.728 | 0.600, 0.884 | 0.175 |
| **Openness to Experience** | 199.581 | < 0.001 | 4.318 | 3.525, 5.290 | 0.806 |

**Table 5:** Odds of challenge volunteer membership by gender, age, education, country of residence and income using logistic regression model, adjusted for HEXACO dimensions.

| Covariate | Category | Wald | p-value | Odds Ratio | Cohen's d |
| --- | --- | --- | --- | --- | --- |
| **Age** | **>43.67 years** | 229.729 | < 0.001 | 0.938 | 0.035 |
| **Country of Residence** | **US Resident** | 84.017 | < 0.001 | 0.032 | 1.898 |
| **Gender** | **Female** | 56.887 | < 0.001 | 0.400 | 0.505 |
|  | **Self-Describe** | 4.032 | 0.045 | 2.234 | 0.443 |
|  | **Prefer not to say** | 1.633 | 0.201 | 0.542 | 0.338 |
| **Education** | **High School** | 9.224 | 0.002 | 26.554 | 1.808 |
|  | **Associate** | 12.235 | < 0.001 | 45.843 | 2.109 |
|  | **Some college** | 18.239 | < 0.001 | 101.516 | 2.547 |
|  | **Bachelor's** | 21.306 | < 0.001 | 144.142 | 2.741 |
|  | **Masters** | 23.931 | < 0.001 | 197.931 | 2.915 |
|  | **Doctoral** | 31.619 | < 0.001 | 526.136 | 3.454 |
|  | **Professional** | 20.433 | < 0.001 | 142.864 | 2.736 |
|  | **Trade/Technical** | 15.812 | < 0.001 | 82.654 | 2.434 |
| **Income** | **\$25k-\$50k** | 9.679 | 0.002 | 1.880 | 0.348 |
| | **\$50k-\$100k** | 28.734 | < 0.001 | 2.873 | 0.582 |
| | **\$100k-\$200k** | 12.297 | < 0.001 | 1.996 | 0.381 |
| | **\$200k +** | 33.161 | < 0.001 | 4.195 | 0.791 |
|  | **Prefer not to say** | 62.390 | < 0.001 | 17.841 | 1.589 |

### Risk sensitivity

We next compared risk behaviors and evaluations across the two groups. We predicted that volunteers would not, in general, exhibit more risk-taking behaviors or risk insensitivity relative to controls^29^. We compared groups on the six DOSPERT risk domains for each of the three components using an ANCOVA model (Table 6 and Supplemental Table 3 and 4), which included an additional covariate for age, and included the categorical variables of income, education level, gender, and US residency as fixed effects to control for the potential role of demographic differences between volunteers and controls. Results indicated that volunteers differed from controls in risk-taking attitudes in all domains. However, the volunteer group was not consistently the more risk-seeking group. Relative to controls, volunteers demonstrated greater risk-aversion in the domains of ethics, gambling, and health and safety. This effect was greatest for risk aversion relating to ethical (V: 1.73, C: 2.60, p<0.001, η^2^=0.113) and financial-gambling scenarios (V: 1.40, C: 2.45, p<0.001,η^2^=0.107). By contrast, volunteers were more risk-seeking than controls with respect to financial investing, recreational activities, and social behaviors (for example, challenging norms or authority). The effect size of risk-seeking was greatest within the social domain (V: 5.39, C: 4.40, p<0.001,η^2^=0.126). Other dimensions of risk-taking showed small differences across the groups (η^2^=0.004-0.028).

**Table 6:**
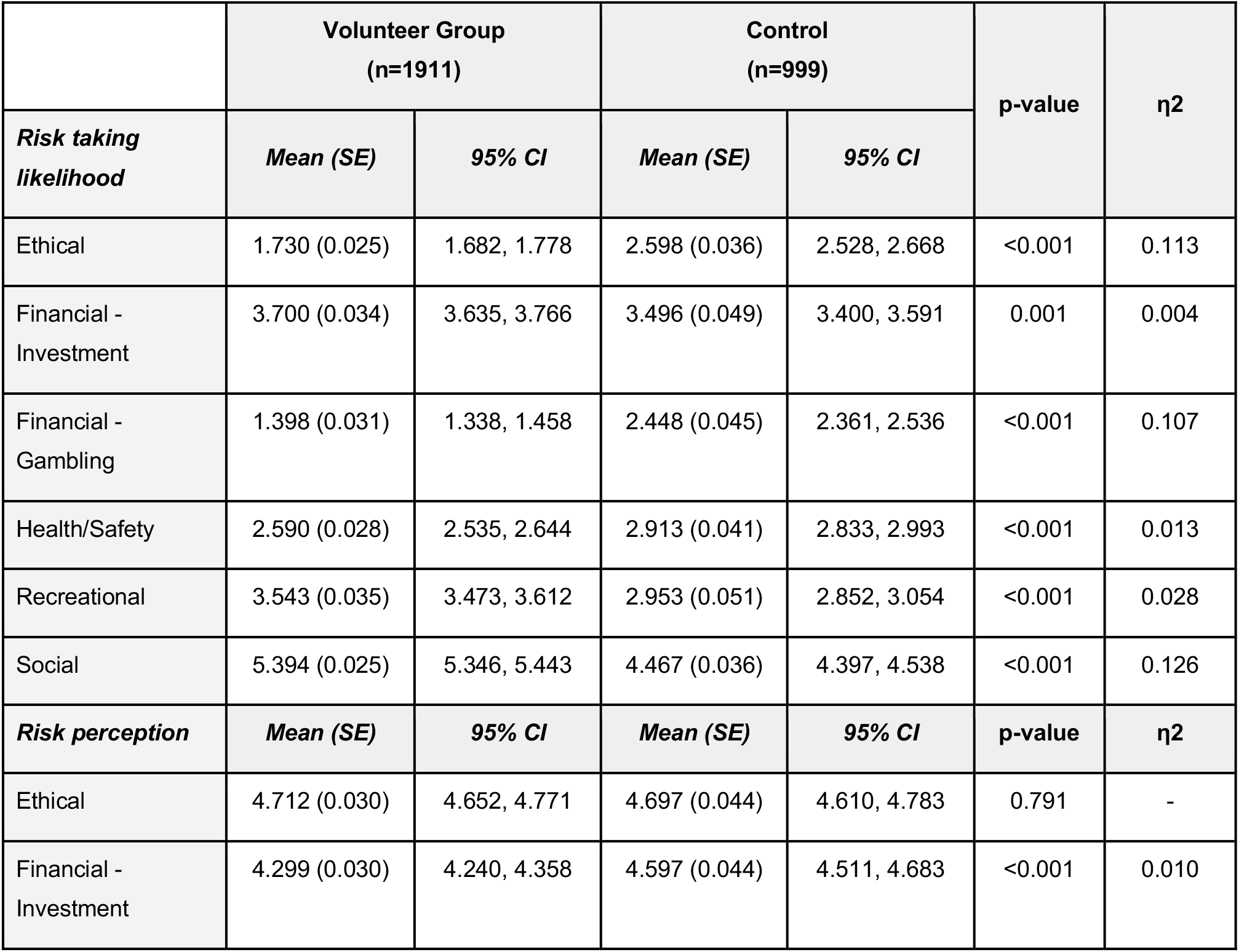
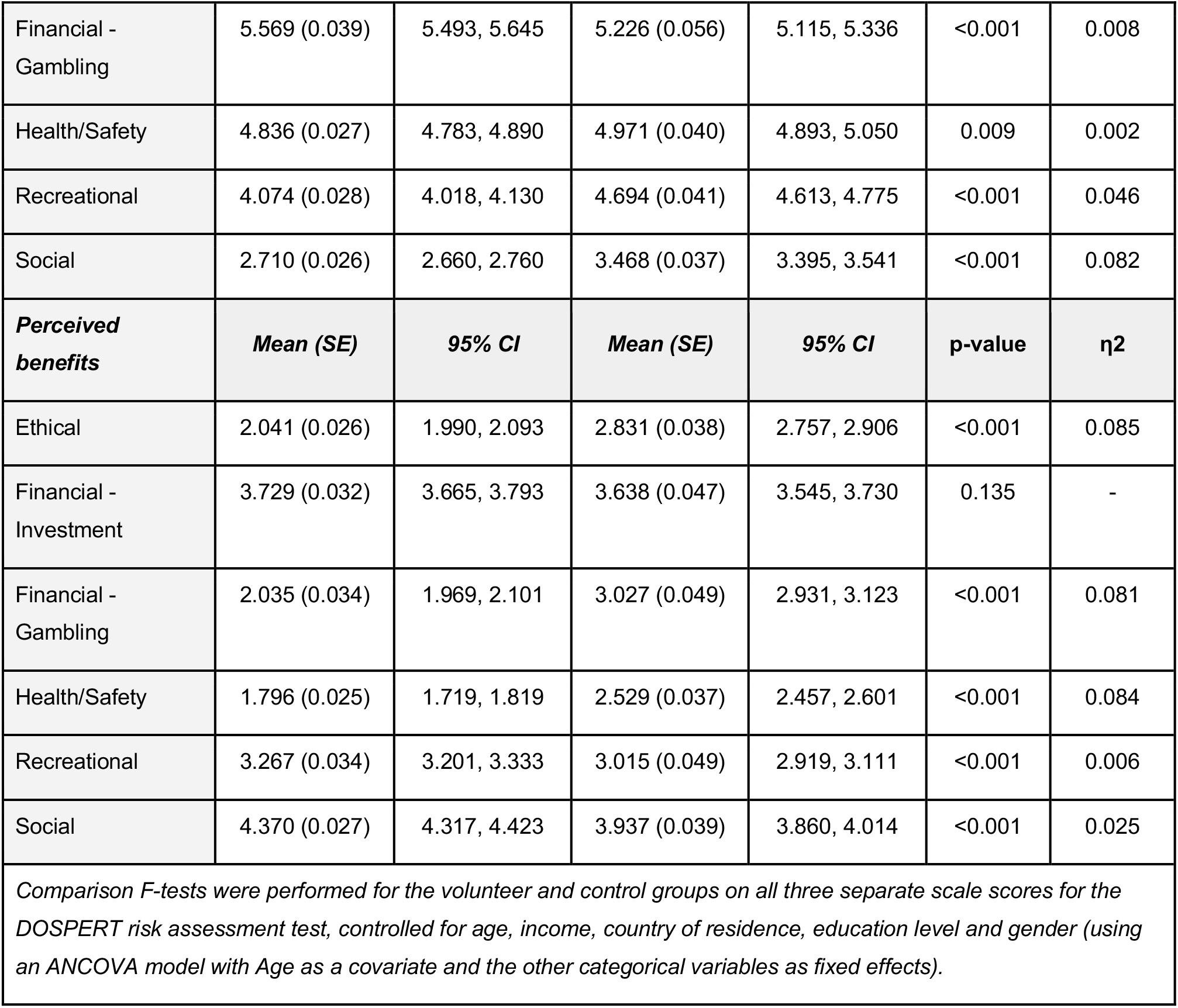
Comparisons of DOSPERT risk attitude component scores.

We also identified significant differences between volunteers and controls on the *risk-perception* component of the DOSPERT across all domains, with the exception of the ethical domain. The strengths of associations were mostly small or trivial (η^2^ <0.01-0.06). However, medium effects were observed for the perception of social risk, which was greater in controls than volunteers (V: 2.71, C: 3.47, p < 0.001, η^2^ = 0.08). Finally, with respect to the *perceived-benefits scale* of the DOSPERT, volunteers perceived risk-taking behaviors in the ethical (V: 2.04, C: 2.83, p < 0.001, η^2^ = 0.085), financial-gambling (V: 2.04, C: 3.03, p < 0.001, η^2^ = 0.081) and health and safety domains (V: 1.80, C: 2.48, p < 0.001, η^2^ = 0.084) as significantly less beneficial than did controls (all medium effect sizes). Exceptions included the recreational (V: 3.27, C: 3.02, p < 0.001,η^2^ = 0.006) and social domains (V: 4.37, C: 3.94, p < 0.001,η^2^ = 0.025), which volunteers perceived as more beneficial than did controls (although differences had trivial and small effect sizes, respectively).

Further analyses regarding risk-perception relating to COVID-19 and challenge trial participation identified in the pre-registration plan were beyond the scope of this paper and will be discussed in forthcoming papers.

## Discussion

Together, these results indicate that the characteristics of volunteers for COVID-19 challenge trials do not substantiate concerns regarding understanding, vulnerability, or undue influence. Volunteerism was overwhelmingly associated with heightened altruistic motivation and behavior. Nearly all volunteers reported altruistic motivations for volunteering, and demonstrated high levels of prior engagement in other forms of altruism, including donating blood, donating money to charity, and registering as living marrow donors and deceased organ donors. Volunteers also scored higher in personality traits like Honesty-Humility that reflect high valuation of others relative to the self^30^. Together, these metrics suggest that those who volunteer to participate in COVID-19 human challenge trials (the benefits of which primarily accrue to others) exhibit reliably altruistic motivations, preferences, and values consistent with the goals of these trials.

We did not find evidence that challenge trial volunteerism is disproportionately associated with psychological or demographic factors that might raise ethical concerns. Comparing risk perceptions and behaviors between volunteers and controls, we found that group differences were generally small in magnitude and did not suggest that volunteers were generally insensitive to factors that compromise physical health or safety. Although volunteers indicated that they would be more likely than controls to take risks in social, recreational, and investment domains, they indicated being less likely to take risks in the health and safety domain. Group differences in ratings may reflect in part the different risk/benefit profiles that the two groups perceived for different categories of risk. Volunteers perceived slightly lower risks in the health and safety domain than controls (η^2^=0.003), but also perceived lower benefits to activities in that domain (η^2^=0.087).

We also found no evidence that volunteerism is associated with high levels of socioeconomic vulnerability that might make volunteers subject to exploitation. Due to the lengthy quarantine period required in COVID-19 challenge trials, the recently approved U.K. trial will compensate volunteers £1,500 for a 17-day quarantine. We cannot rule out the possibility that this payment, meant to compensate for quarantine only, may nonetheless attract people seeking economic gain, which might be construed as coercion or undue inducement to participate (see Largent et al 2017^31^ for a review of the debate on coercion and undue inducement). Our results indicate that challenge trials will likely be able to attract participants with non-economic motives. Volunteers in our sample reported higher levels of income and education relative to population medians and relative to controls, and equivalent levels of health insurance as controls. The high median educational attainment of volunteers (over three-quarters of whom reported having a Bachelor’s degree or higher) also matters, as it suggests that volunteers are relatively well-positioned to understand the information disclosed during the consent process^32^.

Of note, majorities of volunteers were male and between the ages of 18 and 45. A high proportion (78.5%) identified as non-Hispanic white. These socio-demographic variables confer both risk factors for and protective factors against serious COVID-19 outcomes. It is generally accepted that challenge trials should include only young and medically healthy volunteers^2,33^, but the role that other socio-demographic risk factors should play in volunteer enrollment is debated. Male biological sex confers clear risks of serious illness or death following infection, with males’ average case-fatality ratio being 1.7 times higher than females’, an effect thought to reflect sex-based differences in innate and adaptive immune responses^34,35^. COVID-19 related fatalities and hospitalizations are dramatically elevated among participants who identify as Black, Latino, and Native American, likely due to structural inequities and socioeconomic factors affecting health^36^. Some advocates of COVID-19 challenge trials have proposed including volunteers from diverse backgrounds to ensure adequate representation of demographic groups that have been hardest hit by the pandemic^37^. More than twenty percent of the over 38,000 volunteers recruited through 1Day Sooner come from underrepresented groups, suggesting that challenge trials enrolling from this pool could include a diverse group of participants.

Together, these findings are inconsistent with expressed concerns that human challenge trials with the novel coronavirus would be “*prima facie* unethical” because they would be expected to follow a “pattern of exploitative recruitment”^11^. Whereas human challenge trial recruitment could be viewed as inherently exploitative if it attracted volunteers who find participation “very attractive as a result of being in a socioeconomically disadvantaged position as a result of social injustice”^11^ or whose volunteerism reflects “financial desperation, or a distorted understanding of the risks”^38^, our results indicate that such trials tend to attract volunteers who are primarily motivated by altruism and do not on the whole exhibit any indicators of socioeconomic or psychological vulnerability to exploitation.

These results should be interpreted in light of certain limitations. First, the survey was conducted in a sample of early volunteers who signed up with 1Day Sooner in April and May of 2020, the earliest weeks of its creation. Volunteers sampled here may not be representative of all challenge trial volunteers, and those who have subsequently volunteered may be different. We also cannot know what proportion of intended volunteers would pass exclusionary screening and consent to participate in a challenge trial. It is possible that this subset would be small or non-representative of the volunteers characterized in our study, similar to observations that altruistic marrow donors represent only a fraction of those who initially volunteer to donate^39^. However, we have no basis for assuming what specific changes in the composition of challenge trial volunteers would result in. In addition, our sample of controls, whilst recruited to reflect national United States characteristics established by 2019 census data (including age, gender, education and income), are not truly representative of the United States population as a whole. Nor can we rule out, based on our data, the possibility that challenge trial volunteerism reflects unmeasured biases related to the perception of risks and benefits, such as optimism bias^10,40^; the so-called preventative or therapeutic fallacy, which reflects a common assumption that any treatment offered by medical professionals must be potentially beneficial ^41,42^; or unrealistic beliefs about potential personal gains. To some degree, such concerns can be resolved through a robust informed consent process^16,17,43^, which is broadly viewed as possible for COVID-19 challenge trials^2,33,44,45,46^. If, as our findings suggest, intended COVID-19 challenge trial volunteers are mostly aware of and prepared to take the personal risks associated with such studies to benefit the greater good, then, given the large number of intended volunteers to come forth in a short amount of time, we can expect that there will be a sufficient number of altruistic volunteers able to provide valid consent to make these trials both ethical and feasible.

## Conclusions

Self-interest is sometimes incorrectly assumed to be the central or sole value driving human decisions^47,48^, which may contribute to pervasive concerns that volunteerism for risky and primarily other-benefiting biomedical procedures reflects undue inducement or problems understanding consent. However, people vary widely in their selfish versus altruistic preferences and values^30,49^. Those who volunteer for biomedical procedures that confer net personal risks and burdens without direct benefits (like kidney and marrow donations) place unusually high value on others’ welfare relative to their own^39,50^. Such donations are now broadly accepted as ethical despite their risks and absence of direct benefits to volunteers because they are consistent with donors’ values and preferences. In finding that challenge trials can attract volunteers whose altruistic preferences and values align with the goals of these trials (and who are not unusually vulnerable to exploitation), the present findings similarly support the possibility of valid informed consent for COVID-19 challenge trials.

## Methods

### Participants

2,910 individuals completed a 45-minute online survey that included indices of altruistic motivation, values, and behavior; an assessment of risk preferences and behaviors, and a survey of sociodemographic variables. Questions presented to the volunteer and control group are available in Supplementary Data. The sample included 1,911 individuals who had confirmed their willingness to participate in SARS-CoV-2 challenge trials prior to May 29, 2020 and 999 controls. We recruited the maximum sample size possible within our financial constraints. Potential challenge trial volunteers were recruited through the non-profit advocacy organization 1Day Sooner. Volunteers who had declared their intent to volunteer and provided their contact information as well as their interest in participating in research were recruited via email (Figure 2).

**Figure 2:**
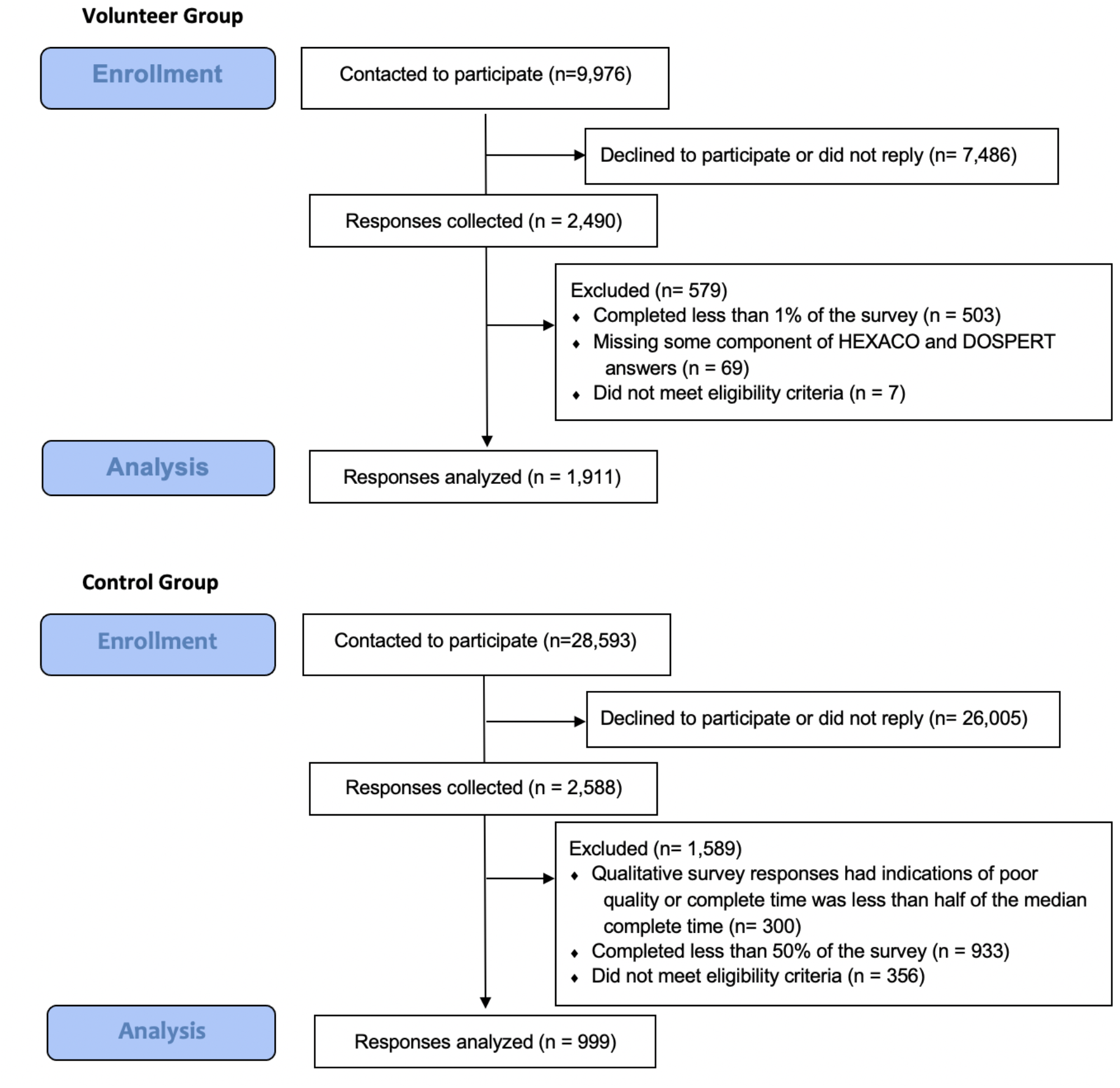
CONSORT Diagram of Volunteer and Control Group Enrollment and Analysis.

Control participants were recruited using a private research software company (Qualtrics Panel), which identifies individuals through other survey-hosting platforms and is intended to be reflective of the population distribution captured by the 2019 United States Census. Inclusion criteria for all participants included age greater than 18 years and demonstrated proficiency in English. Volunteers in the database were excluded from sampling if they were under age 18, responded ‘no’ to a query about wanting to participate in a vaccine challenge trial, declined to share their information with researchers, or declined to provide a response to a query about reasons for participating (open response format). Participants who responded to this question were filtered if they responded in a language other than English, or if responses were too brief (<5 words) to ascertain fluency in English. All participants who completed the survey were compensated $5 USD in the form of an electronic gift card. Participants who expressed interest in completing the survey were allotted 7 days to complete it at a time of their choosing, and could complete it in more than one sitting if they preferred. Those who did not complete the survey were sent follow-up emails on Day 4 and Day 6 to give them the opportunity to complete their response. The protocol was approved by the Institutional Review Board at Rutgers University (Study ID: Pro2020001023) and all participants provided electronic informed consent before beginning the survey. All statistical tests for this study were taken from the same sample and are two-tailed tests.

### Survey Instruments

Indices of altruistic values and preferences were as follows: First, the volunteer group selected their top three motivations for volunteering from a list of 10 possible motivations drawn from consultations with a panel of challenge study researchers and bioethicists (Table 2) (controls did not complete this section)^16,18,25,51^. Two motivations were primarily altruistic in that they refer to outcomes for entities outside the self (“I wanted to help others and potentially save lives” and “I wanted to contribute to the progress of medicine”); the other 8 reflected various other motivations (e.g., “I wanted to receive the financial reimbursement for participating” or “I was curious about COVID-19”). Second, participants indicated their prior engagement in various altruistic behaviors that carry varying levels of risk and cost, including blood donation, registering to donate bone marrow, registering to be a deceased organ donor, donating money to charity, and living organ donation. Participants completed two additional instruments assessing personality traits and risk perception. The Brief HEXACO inventory is a 24-item measure assessing six dimensions of personality: Honesty-Humility, Emotionality, eXtraversion, Agreeableness, Conscientiousness, and Openness to Experience^52,53^. Each item is rated on a five-point scale. Unlike five-factor inventories, HEXACO inventories include a subscale (Honesty-Humility) that specifically indexes attitudes and behaviors related to valuation of outcomes for others versus the self (such as exploitation, manipulation, or deceit) and has been consistently linked to prosocial motivation and behavior^30,54,55^. The DOSPERT scale is a 30-item index that assesses three primary components of risk attitudes (risk-taking, risk-perception and perceived expected benefits) across six broad decision categories: ethical, financial (divided into investment and gambling), health and safety, social, and recreational risks^56^. The *risk-taking scale* assesses respondents’ likelihood of engaging in the risky activity or behavior, the *risk-perception scale* assesses how risky participants perceive each of these activities to be, and the *expected-benefits scale* assesses how beneficial participants perceive each activity to be. Responses are made using a 7-point scale (1 = Extremely unlikely/Not at all risky/No benefits at all, 7 = Extremely likely/ Extremely risky/Great benefits). Finally, all participants completed an assessment of socioeconomic and other demographic variables (see Table 1 for a description of these demographic characteristics). Regression models for all analyses included the covariates of age, gender, education level, income and country of residence to control for the potential influence of differences in these characteristics. Age was included as a continuous (scale) variable, centered at the mean age of 43.67 years. Gender was analyzed as a categorical variable, broken down into male (reference), female, self-describe or prefer not to say. Education was analyzed as a categorical variable (high school equivalent or less, trade or technical school, associate degree, some college, bachelor’s degree, masters degree, professional or doctoral degree). Income was analyzed as a categorical variable with six categories: less than $25k annual household income, $25k-$50k, $50k-$100k, $100k-$200k, greater than $200k, and prefer not to say. Country of residence was dichotomized as non-US (reference) and US.

### HEXACO Analysis

Analysis of the HEXACO results began with an exploratory factor analysis (EFA) to assess factor components. The EFA found six factors that roughly corresponded to the six dimensions captured by the HEXACO model (r^2^=0.526) (see Supplementary Methods and Supplementary Table 2 for a comparison of the HEXACO dimensions and Survey EFA components by question). Therefore, subsequent analyses were performed using the standard six HEXACO dimensions. Firstly, scores on each HEXACO dimension were compared across volunteer and control groups using an ANCOVA model controlling for age, income, education level, country of residence and gender. Subsequent analyses were conducted to determine the likelihood of participants being in the volunteer or control group based on their HEXACO scores. These analyses were initially conducted using a multivariate logistic regression containing only the six HEXACO dimensions as independent variables, and membership in the volunteer group as the dependent variable. An additional multivariate analysis including the demographic covariates of age, gender, education level, income, and US residency was then conducted.

### DOSPERT Analysis

Preliminary analysis of the DOSPERT was also performed using EFA to match the factors of the two groups to the DOSPERT domains. A separate EFA was performed on each component of the DOSPERT scale (risk-taking, risk perception, and perceived expected benefits) (See Supplementary Methods for a comparison of the DOSPERT factors across the three components). As the association between the DOSPERT component and EFA results were moderate for all three components (r^2^=0.536, 0.568 and 0.614 respectively), further analyses were performed using the original six DOSPERT domains: Ethical, Financial - Investment, Financial - Gambling, Health/Safety, Recreational and Social. Risk behaviors and evaluations were compared across the volunteer and control groups using an ANCOVA model that included an additional covariate for age, and included the categorical variables of income, education level, gender, and country of residence as fixed effects to control for the potential role of demographic differences between volunteers and controls.

## Data Availability

The anonymized data set was uploaded to a github repository and the doi is as follows: 10.5281/zenodo.4601486

https://doi.org/10.5281/zenodo.4601486

## Figure Captions

***Figure 1*. (A)** Participants in the volunteer group were asked to indicate their top three motivations for participating in a COVID-19 challenge trial from a list of ten options. Selections were not ranked, and total percentages add to 300% because each participant selected 3 options. The two most commonly selected options were “I wanted to help others and potentially save lives” (95.9%) and “I wanted to contribute to the progress of medicine” (79.2%). **(B)** Participants in volunteer and control groups were surveyed on their engagement with a range of altruistic behaviors, including blood donation, significant charitable donations and organ/marrow donor status. Volunteers were significantly more likely than controls to have participated in all but one of the altruistic behaviors.

***Figure 2***. Overall, 9,976 volunteers from the 1Day Sooner database who had indicated they were interested in contributing to further research were contacted to participate in our study. Of these, 7,486 volunteers did not reply or declined to participate. The remaining 2,490 volunteers completed the survey via the Qualtrics platform. 579 of these responses were ultimately excluded from the final analysis, due to failure to complete sufficient portions of the survey, missing data or submitting a birth date that indicated they were under 18 years of age. The remaining 1,911 responses were then analyzed.

## References

1. Johnson, C. Y. Variants mean the coronavirus is here to stay — but perhaps as a lesser threat. Washington Post (2021).

2. World Health Organization. Key criteria for the ethical acceptability of COVID-19 human challenge studies. (2020).

3. Meiring, J. E., Giubilini, A., Savulescu, J., Pitzer, V. E. & Pollard, A. J. Generating the Evidence for Typhoid Vaccine Introduction: Considerations for Global Disease Burden Estimates and Vaccine Testing Through Human Challenge. Clinical Infectious Diseases 69, S402–S407 (2019).

4. Cooper, M. M., Loiseau, C., McCarthy, J. S. & Doolan, D. L. Human challenge models: tools to accelerate the development of malaria vaccines. Expert Review of Vaccines 18, 241–251 (2019).

5. Roestenberg, M., Hoogerwerf, M.-A., Ferreira, D. M., Mordmüller, B. & Yazdanbakhsh, M. Experimental infection of human volunteers. The Lancet Infectious Diseases 18, e312–e322 (2018).

6. Roestenberg, M., Kamerling, I. M. C. & de Visser, S. J. Controlled Human Infections As a Tool to Reduce Uncertainty in Clinical Vaccine Development. Front. Med. 5, (2018).

7. Sauerwein, R. W., Roestenberg, M. & Moorthy, V. S. Experimental human challenge infections can accelerate clinical malaria vaccine development. Nature Reviews Immunology 11, 57–64 (2011).

8. Hodgson, S. H. et al. What defines an efficacious COVID-19 vaccine? A review of the challenges assessing the clinical efficacy of vaccines against SARS-CoV-2. The Lancet Infectious Diseases 21, e26–e35 (2021).

9. Nguyen, L. C. et al. Evaluating Use Cases for Human Challenge Trials in Accelerating SARS-CoV-2 Vaccine Development. Clinical Infectious Diseases 72, 710–715 (2021).

10. Kahn, J. P., Henry, L. M., Mastroianni, A. C., Chen, W. H. & Macklin, R. Opinion: For now, it’s unethical to use human challenge studies for SARS-CoV-2 vaccine development. PNAS 117, 28538–28542 (2020).

11. Holm, S. Controlled human infection with SARS-CoV-2 to study COVID-19 vaccines and treatments: bioethics in Utopia. Journal of Medical Ethics 46, 569–573 (2020).

12. Bramble, B. Challenge trials for a coronavirus vaccine are unethical – except for in one unlikely scenario. The Conversation (2020).

13. Franklin, G. M. & Grady, C. The Ethical Challenge of Infection-Inducing Challenge Experiments. Clinical Infectious Diseases 33, 1028–1033 (2001).

14. Evers, D. L., Fowler, C. B., Mason, J. T. & Mimnall, R. K. Deliberate Microbial Infection Research Reveals Limitations to Current Safety Protections of Healthy Human Subjects. Sci Eng Ethics 21, 1049–1064 (2015).

15. Department for Business, Energy & Industrial Strategy & The Rt Hon Kwasi Kwarteng MP. World’s first coronavirus Human Challenge study receives ethics approval in the UK. GOV.UK https://www.gov.uk/government/news/worlds-first-coronavirus-human-challenge-study-receives-ethics-approval-in-the-uk (2021).

16. Njue, M. et al. Ethical considerations in Controlled Human Malaria Infection studies in low resource settings: Experiences and perceptions of study participants in a malaria Challenge study in Kenya [version 2; peer review: 2 approved]. Wellcome Open Res 3, (2018).

17. Kraft, S. A. et al. Exploring Ethical Concerns About Human Challenge Studies: A Qualitative Study of Controlled Human Malaria Infection Study Participants’ Motivations and Attitudes. Journal of Empirical Research on Human Research Ethics 14, 49–60 (2019).

18. Jao, I. et al. Deliberately infecting healthy volunteers with malaria parasites: Perceptions and experiences of participants and other stakeholders in a Kenyan-based malaria infection study. Bioethics 34, 819–832 (2020).

19. Miller, F. G. The ethical challenges of human research: selected essays. (Oxford University Press, 2012).

20. National Commission for the Protection of Human Subjects of Biomedical and Behavioral Research. The Belmont Report. https://www.hhs.gov/ohrp/regulations-and-policy/belmont-report/read-the-belmont-report/index.html (1979).

21. Schenker, Y. & Meisel, A. Informed Consent in Clinical Care: Practical Considerations in the Effort to Achieve Ethical Goals. JAMA 305, 1130–1131 (2011).

22. Evans, D. An activist’s argument that participant values should guide risk–benefit ratio calculations in HIV cure research. Journal of Medical Ethics 43, 100–103 (2017).

23. Sylla, L. et al. If We Build It, Will They Come? Perceptions of HIV Cure-Related Research by People Living with HIV in Four U.S. Cities: A Qualitative Focus Group Study. AIDS Research and Human Retroviruses 34, 56–66 (2018).

24. Murray, B. R. et al. What risk of death would people take to be cured of HIV and why? A survey of people living with HIV. Journal of Virus Eradication 5, 109–115 (2019).

25. Oguti, B. et al. Factors influencing participation in controlled human infection models: a pooled analysis from six enteric fever studies [version 1; peer review: 3 approved with reservations]. Wellcome Open Res 4, 153 (2019).

26. Semega, J., Kollar, M., Shrider, E. A. & Creamer, J. Income and Poverty in the United States: 2019. https://www.census.gov/library/publications/2020/demo/p60-270.html (2020).

27. Cohen, J. Statistical power analysis for the behavioral sciences. (L. Erlbaum Associates, 1988).

28. McElreath, R. Statistical rethinking: a Bayesian course with examples in R and Stan. (CRC Press, 2018).

29. Marsh, A. A. et al. Neural and cognitive characteristics of extraordinary altruists. PNAS 111, 15036–15041 (2014).

30. Zettler, I., Thielmann, I., Hilbig, B. E. & Moshagen, M. The Nomological Net of the HEXACO Model of Personality: A Large-Scale Meta-Analytic Investigation. Perspect Psychol Sci 15, 723–760 (2020).

31. Largent, E. A. & Lynch, H. F. Paying Research Participants: Regulatory Uncertainty, Conceptual Confusion, and a Path Forward. Yale J Health Policy Law Ethics 17, 61–141 (2017).

32. Christopher, P. P., Foti, M. E., Roy-Bujnowski, K. & Appelbaum, P. S. Consent Form Readability and Educational Levels of Potential Participants in Mental Health Research. PS 58, 227–232 (2007).

33. Eyal, N., Lipsitch, M. & Smith, P. G. Human Challenge Studies to Accelerate Coronavirus Vaccine Licensure. The Journal of Infectious Diseases 221, 1752–1756 (2020).

34. Scully, E. P., Haverfield, J., Ursin, R. L., Tannenbaum, C. & Klein, S. L. Considering how biological sex impacts immune responses and COVID-19 outcomes. Nature Reviews Immunology 20, 442–447 (2020).

35. Peckham, H. et al. Male sex identified by global COVID-19 meta-analysis as a risk factor for death and ITU admission. Nature Communications 11, (2020).

36. Rossen, L. M., Branum, A. M., Ahmad, F. B., Sutton, P. & Anderson, R. N. Excess Deaths Associated with COVID-19, by Age and Race and Ethnicity — United States, January 26– October 3, 2020. MMWR Morb Mortal Wkly Rep 69, 1522–1527 (2020).

37. McPartlin, S. O., Morrison, J., Rohrig, A. & Weijer, C. Covid-19 vaccines: Should we allow human challenge studies to infect healthy volunteers with SARS-CoV-2? BMJ 371, m4258 (2020).

38. Wolemonwu, V. C. Human Challenge Trials for a COVID-19 Vaccine: Should we bother about exploitation? VIB 6, (2020).

39. Vekaria, K. M. et al. The role of prospection in altruistic bone marrow donation decisions. Health Psychology 39, 316–324 (2020).

40. Weinstein, N. D. Unrealistic optimism about future life events. Journal of Personality and Social Psychology 39, 806–820 (1980).

41. Jansen, L. A. et al. Unrealistic Optimism in Early-Phase Oncology Trials. IRB 33, 1–8 (2011).

42. Horng, S. & Grady, C. Misunderstanding in Clinical Research: Distinguishing Therapeutic Misconception, Therapeutic Misestimation, & Therapeutic Optimism. IRB: Ethics & Human Research 25, 11–16 (2003).

43. Cassileth, B. R., Zupkis, R. V., Sutton-Smith, K. & March, V. Informed Consent — Why Are Its Goals Imperfectly Realized? N Engl J Med 302, 896–900 (1980).

44. Jamrozik, E. & Selgelid, M. J. Ethical Issues. in Human Challenge Studies in Endemic Settings: Ethical and Regulatory Issues 25–82 (Springer International Publishing, 2021). doi:10.1007/978-3-030-41480-1_3.

45. Chappell, R. Y. & Singer, P. Pandemic ethics: the case for risky research. Research Ethics 16, 1–8 (2020).

46. WHO Expert Committee on Biological Standardization. Human Challenge Trials for Vaccine Development: regulatory considerations. (2016).

47. Miller, D. T. The norm of self-interest. American Psychologist 54, 1053–1060 (1999).

48. Elster, J. Chapter 3 Altruistic Behavior and Altruistic Motivations. in Handbook of the Economics of Giving, Altruism and Reciprocity (eds. Kolm, S.-C. & Ythier J. M.) vol. 1 183– 206 (Elsevier, 2006).

49. Schwartz, S. H. Universals in the Content and Structure of Values: Theoretical Advances and Empirical Tests in 20 Countries. in Advances in Experimental Social Psychology (ed. Zanna, M. P.) vol. 25 1–65 (Academic Press, 1992).

50. Vekaria, K. M., Brethel-Haurwitz, K. M., Cardinale, E. M., Stoycos, S. A. & Marsh, A. A. Social discounting and distance perceptions in costly altruism. Nature Human Behaviour 1, 1–7 (2017).

51. Hoogerwerf, M.-A., Vries M. de & Roestenberg, M. Money-oriented risk-takers or deliberate decision-makers: a cross-sectional survey study of participants in controlled human infection trials. BMJ Open 10, e033796 (2020).

52. de Vries, R. E. The 24-item Brief HEXACO Inventory (BHI). Journal of Research in Personality 47, 871–880 (2013).

53. Lee, K. & Ashton, M. C. Psychometric Properties of the HEXACO Personality Inventory. Multivariate Behavioral Research 39, 329–358 (2004).

54. Brocklebank, S., Pauls, S., Rockmore, D. & Bates, T. C. A spectral clustering approach to the structure of personality: Contrasting the FFM and HEXACO models. Journal of Research in Personality 57, 100–109 (2015).

55. Weller, J. A. & Tikir, A. Predicting domain-specific risk taking with the HEXACO personality structure. Journal of Behavioral Decision Making 24, 180–201 (2011).

56. Blais, A.-R. & Weber, E. U. A Domain-Specific Risk-Taking (DOSPERT) Scale for Adult Populations. Judgment and Decision Making 1, 33–47 (2006).

