## Supplemental data for "Characterizing altruistic motivation in potential volunteers for SARS-CoV-2 challenge trials"

**Supplementary Tables**

This section consists of Supplementary Tables 1 - 5.

**Supplementary Table 1: Factor Loading Table for HEXACO:**

| Question | Factor 1 (H) | Factor 2 (O) | Factor 3 (C) | Factor 4 (X) | Factor 5 (E) | Factor 6 (A) |
|---|---|---|---|---|---|---|
| 6 | .065 | .062 | .462 | .089 | .149 | .341 |
| 12R | .714 | -.023 | .120 | .014 | .125 | .090 |
| 18R | .590 | -.219 | .016 | -.099 | .031 | -.003 |
| 24R | .712 | .101 | -.064 | -.064 | -.023 | .036 |
| 5 | -.413 | -.093 | .145 | .015 | .487 | -.030 |
| 11R | .180 | .076 | -.030 | -.303 | .664 | -.116 |
| 17R | -.025 | -.207 | -.384 | -.206 | .536 | -.007 |
| 23 | -.254 | .063 | .099 | .308 | .494 | .183 |
| 4R | .665 | .002 | -.005 | .368 | -.013 | .081 |
| 10 | -.058 | .146 | .090 | .762 | -.225 | -.008 |
| 16 | .040 | .226 | .106 | .778 | .023 | .032 |
| 22R | .580 | -.063 | -.047 | .439 | -.072 | .152 |
| 3R | .362 | .152 | -.221 | .094 | -.190 | .539 |
| 9R | .237 | -.250 | -.027 | -.034 | -.033 | .688 |
| 15 | -.411 | .154 | .139 | .046 | .176 | .416 |
| 21 | -.070 | .311 | .147 | .084 | -.461 | .465 |
| 2 | .007 | .059 | .744 | .084 | .008 | -.047 |
| 8R | .497 | -.078 | .272 | .023 | -.310 | .098 |
| 14 | .079 | .167 | .754 | .015 | -.154 | -.082 |
| 20R | .608 | -.118 | .256 | -.102 | -.118 | .064 |
| 1 | -.019 | .566 | .253 | .090 | .057 | .093 |
| 7R | .630 | .399 | -.080 | -.076 | -.027 | .000 |
| 13 | .032 | .654 | .170 | .190 | -.023 | -.086 |

| | | | | | | |
|---|---|---|---|---|---|---|
| 19 | -.104 | .757 | -.081 | .071 | -.118 | .010 |

**Supplementary table 1:** After controlling for gender, education level, income, and country of residence between volunteers and controls, this table demonstrates that volunteer status was most strongly associated with Openness to experience and Honesty-Humility while Agreeableness was not associated more strongly with either volunteer or control respondents. The column on the far left is the question number that was presented to those responding to the questionnaire and the top row illustrates which factor from the HEXACO paradigm is represented by each factor. The highest loaded factor is highlighted, and the closest HEXACO factor given in parentheses.

**Supplementary table 2: Exploratory Factor Analysis Summary of Survey Components compared to HEXACO Factors**

| HEXACO Factors | Survey EFA Components |
|---|---|
| Honesty/Humility: Questions 6, 12R, 18R, 24R | Mostly Honesty/Humility & reverse-codes: Questions 4R, 7R, 8R, 12R, 18R, 20R, 22R, 24R |
| Emotionality: Questions 5, 11R, 17R, 23 | Emotionality: Questions 5, 11R, 17R, 23 |
| eXtraversion: Questions 4R, 10, 16, 22R | Mostly eXtraversion: Questions 10, 16 |
| Agreeableness: Questions 3R, 9R, 15, 21 | Agreeableness: Questions 3R, 9R, 15, 21 |
| Conscientiousness: Questions 2, 8R, 14, 20R | Mostly Conscientiousness + Question 6: Questions 2, 6, 14 |
| Openness to Experience: Questions 1, 7R, 13, 19 | Mostly Openness to Experience: Questions 1, 13, 19 |

**Supplementary table 2:** Results from these analyses indicate that Honesty/Humility and Openness to Experience are the HEXACO scale scores that are the most indicative to volunteer group membership, with most effect sizes in the medium-to-large range. Most of the other factors were either insignificant or had a trivial or near trivial effect.

**Supplementary Table 3: Exploratory Factor Analysis Summary of Survey Components compared to DOSPERT Scale Components**

| **DOSPERT Scale Factor** | **DOSPERT Scale Component** | | |
|---|---|---|---|
| | ***Risk-Taking Likelihood EFA*** | ***Risk Perception EFA*** | ***Perceived Benefits EFA*** |
| Ethical (6,9,10,16,29,30) | Ethical & Financial-Gambling (3,6,8,10,14) | Ethical & Health/Safety (5,6,9,10,15,16,17,20,23,29,30) | Ethical & Health/Safety (5,6,9,10,15,16,17,20,23,26,29,30) |
| Financial-Investing (4,12,18) | Financial-Investing (4,12,18) | Financial-Investing & Financial-Gambling (3,8,12,14,18) | Financial-Investing & Financial-Gambling (3,4,8,12,14,18) |
| Financial-Gambling (3,8,14) | | | |
| Health/Safety (5,15,17,20,23,26) | Health/Safety + remainder of Ethical (5,9,15,16,17,20,23, 26,29,30) | | |
| Recreational (2,11,13,19,24,25) | Recreational (2,11,13,19,24,25) | Recreational (11,13,19,24,25,26) | Recreational (2,11,13,19,24,25) |
| Social (1,7,21,22,27,28) | Social (1,7,21,22,27,28) | Social (1,7,21,22,27,28) | Social 1 (21,27,28) |
| | | Remainder (2,4) | Social 2 (1,7,22) |

**Supplementary table 3:** For the EFA results, factor names are derived from the closest DOSPERT factor(s) that match the questions present for that factor, with “Remainder” used for the last (lowest eigenvalued) factor in the risk perception category that did not closely correspond to any DOSPERT factor(s).

Results of the EFA on risk-taking likelihood gave a five factor solution, explaining 53.6% of the variance, with the main difference being that the Ethical domain questions were divided between the Financial-Gambling and Health/Safety domains

Results of the EFA on risk perception also gave a five factor solution, explaining 56.8% of the variance, with most of the questions from the Financial-Gambling and Financial-Investment domains combined and most of the questions in the Ethical and Health/Safety domains combined, one question and a small, two-question Remainder domain (one from Financial-Investing and one from Recreational) as the fifth factor with a marginal eigenvalue of 1.2

Results of the EFA on perceived benefits also gave a five factor solution, explaining 61.4% of the variance, with the Financial-Investment and Financial-Gambling domains combined, Ethical and Health/Safety domains combined, and the Social domain split into two factors (with three questions each).

**Supplementary Table 4: Comparisons of DOSPERT risk attitude component scores**

| | Volunteer Group (n=1911) | | Control (n=999) | | p-value | η2 |
|---|---|---|---|---|---|---|
| ***Risk taking likelihood*** | ***Mean (SE)*** | ***95% CI*** | ***Mean (SE)*** | ***95% CI*** | | |
| Ethical | 2.082 (0.167) | 1.754, 2.410 | 2.941 (0.168) | 2.611, 3.270 | <0.001 | 0.115 |
| Financial - Investment | 3.711 (0.227) | 3.265, 4.157 | 3.505 (0.228) | 3.057, 3.953 | 0.001 | 0.004 |
| Financial - Gambling | 1.739 (0.208) | 1.332, 2.147 | 2.776 (0.209) | 2.367, 3.185 | <0.001 | 0.109 |
| Health/Safety | 2.773 (0.191) | 2.399, 3.146 | 3.084 (0.191) | 2.709, 3.459 | <0.001 | 0.013 |
| Recreational | 3.816 (0.240) | 3.346, 4.286 | 3.217 (0.241) | 2.745, 3.689 | <0.001 | 0.030 |
| Social | 5.268 (0.167) | 4.940, 5.595 | 4.351 (0.168) | 4.022, 4.680 | <0.001 | 0.129 |
| ***Risk perception*** | ***Mean (SE)*** | ***95% CI*** | ***Mean (SE)*** | ***95% CI*** | **p-value** | **η2** |
| Ethical | 4.581 (0.205) | 4.178, 4.984 | 4.589 (0.206) | 4.185, 4.994 | 0.884 | - |
| Financial - Investment | 4.077 (0.204) | 3.677, 4.477 | 4.370 (0.205) | 3.968, 4.772 | <0.001 | 0.010 |
| Financial - Gambling | 5.300 (0.262) | 4.786, 5.815 | 4.987 (0.263) | 4.471, 5.504 | <0.001 | 0.007 |
| Health/Safety | 4.801 (0.186) | 4.436, 5.165 | 4.945 (0.187) | 4.579, 5.312 | 0.004 | 0.003 |
| Recreational | 3.961 (0.193) | 3.582, 4.340 | 4.605 (0.194) | 4.225, 4.985 | <0.001 | 0.052 |
| Social | 2.849 (0.174) | 2.508, 3.189 | 3.632 (0.174) | 3.290, 3.974 | <0.001 | 0.091 |
| ***Perceived benefits*** | ***Mean (SE)*** | ***95% CI*** | ***Mean (SE)*** | ***95% CI*** | **p-value** | **η2** |
| Ethical | 2.262 (0.178) | 1.914, 2.611 | 3.051 (0.179) | 2.700, 3.401 | <0.001 | 0.088 |
| Financial - Investment | 3.624 (0.220) | 3.193, 4.055 | 3.541 (0.221) | 3.108, 3.974 | 0.161 | - |
| Financial - Gambling | 2.051 (0.229) | 1.603, 2.500 | 3.033 (0.230) | 2.583, 3.484 | <0.001 | 0.083 |
| Health/Safety | 1.967 (0.172) | 1.629, 2.305 | 2.727 (0.173) | 2.387, 3.066 | <0.001 | 0.087 |
| Recreational | 3.288 (0.228) | 2.841, 3.735 | 3.036 (0.229) | 2.587, 3.485 | <0.001 | 0.006 |
| Social | 4.402 (0.183) | 4.042, 4.761 | 3.972 (0.184) | 3.611, 4.333 | <0.001 | 0.026 |

*Comparison t-tests were performed for the volunteer and control groups on all three separate scale scores for the DOSPERT risk assessment test, controlled for age, income, education level and gender (using an ANCOVA model with Age as a covariate and the other categorical variables as fixed effects).*

**Supplementary table 4:** Covariates for age were included in this analysis. The categorical variable of income, education level, gender, and US residency were fixed effects to control for the potential role of demographic differences between volunteer and control respondents. This table looks at the DOSPERT risk assignments in bold and further breaks down the areas of interest in that test analyzed by various questions in the questionnaire posed to all of the respondents. Relative to controls, volunteers demonstrated greater risk aversion in the domains of ethics, gambling, and health and safety. While volunteers were more risk seeking than controls with respect to financial investing, recreational activities, and social behaviors. Other dimensions of risk-taking showed small differences across the groups.

#### Supplementary Methods

This section presents additional data along with more detailed descriptions of the analyses performed on the data provided by the volunteer and control group respondents for the HEXACO analyses as well as the motivation questions broken out for the EFA analysis. Further, a discussion of EFA analyses is presented. Finally, the complete list of study questions presented to the volunteer and control respondents are also presented in this section.

*Volunteer/Control Group Membership and HEXACO Scales*

An analysis was conducted to determine the likelihood of participants being in the volunteer or control group based on their HEXACO scores. This was done using a logistic regression model, with presence in the volunteer/control group as the dependent variable, and the HEXACO scores as the independent variables. This model had an r-squared (Cox & Snell) of 0.246, with the results of the HEXACO scalpel scores given in the table below. OR refers to the odds ratio, and Cohen's d is calculated from the odds ratio using the formula (Hasselblad & Hedges, 1995, Chinn, 2000):

$$ln(OR) \times \frac{\sqrt{3}}{\pi}$$

Standard levels for Cohen's d are given below (Cohen, 1988):

Trivial effect: Less than 0.2
Small effect: 0.2 - 0.5
Medium effect: 0.5 - 0.8
Large effect: Greater than 0.8

To account for possible differences based on age, gender, education level, and income, a second logistic model was performed with those variables as covariates. This model had a total Cox & Snell r-squared of 0.397 (with the HEXACO scale scores block accounting for 0.178 of that r-squared) with the results of the HEXACO variables for this model given in the table below:

**<u>Motivation Question HEXACO EFA</u>**: The table presenting the data obtained from this methodology is presented in **Supplemental Table 2**.

An exploratory factor analysis was performed on the responses of HCT volunteers to the following questions regarding their motivations for volunteering:

Question 2.6_1: I want to help others and potentially save lives

Question 2.6_2: I feel helpless and this is a way to do something positive

Question 2.6_3: I was curious about COVID-19

Question 2.6_4: I wanted to be part of a clinical trial

Question 2.6_5: I wanted to contribute to the progress of medicine

Question 2.6_6: I wanted to find out more about my own health

Question 2.6_7: I wanted to receive financial reimbursement for participating

Question 2.6_8: I wanted to be guaranteed access to critical care should I be infected with COVID-19

Question 2.6_9: I am likely to be infected with COVID-19 anywayThe EFA analysis gave a 3 factor solution (variance explained 60.2%), with results given belowRotated Component Matrix (Varimax Rotation):

| Question number | Factor 1 | Factor 2 | Factor 3 |
|---|---|---|---|
| Question 2.6_1 | -0.478 | 0.374 | 0.534 |
| Question 2.6_2 | 0.174 | 0.003 | 0.832 |
| Question 2.6_3 | 0.273 | 0.688 | 0.026 |
| Question 2.6_4 | 0.163 | 0.790 | -0.069 |
| Question 2.6_5 | -0.253 | 0.730 | 0.236 |
| Question 2.6_6 | 0.587 | 0.522 | -0.022 |
| Question 2.6_7 | 0.722 | 0.121 | -0.199 |
| Question 2.6_8 | 0.713 | 0.200 | 0.132 |
| Question 2.6_9 | 0.574 | -0.105 | 0.289 |

Resulting factors (with suggested titles from the author):

**Factor 1 (Personal Benefit):**

Question 2.6_6: I wanted to find out more about my own health

Question 2.6_7: I wanted to receive financial reimbursement for participating

Question 2.6_8: I wanted to be guaranteed access to critical care should I be infected with COVID-19

Question 2.6_9: I am likely to be infected with COVID-19 anyway

**Factor 2 (Curiosity/Science):**

Question 2.6_3: I was curious about COVID-19

Question 2.6_4: I wanted to be part of a clinical trial

Question 2.6_5: I wanted to contribute to the progress of medicine

**Factor 3 (Altruism):**

Question 2.6_1: I want to help others and potentially save lives

Question 2.6_2: I feel helpless and this is a way to do something positive

Note: Question 2.6_6 (I want to find out more about my own health) had a loading of 0.587 into Factor 1, but also had a loading of 0.522 into Factor 2, so it might belong in Factor 2.

**Survey questions:**
Below are the questions that were sent to each participant

Information that was provided to participants at the beginning of the survey:

This survey will not be used to assess eligibility or identify potential participants for any future human challenge trial studies. Your answers will remain anonymous, and have no bearing on your participation in further research, and will not influence your ability to participate in future research in any way.

A COVID-19 infection study is likely to span a couple of months, with at least several weeks spent isolated in a dedicated clinical facility. Initially, volunteers would be screened for infection and undergo antibody testing to ensure they have not been exposed to the virus. Subsequently, participants would receive either placebo or vaccine. The virus would be administered roughly 2-4 weeks later after participants have had time to mount an immune response to the vaccine, after which participants would be observed from the time of infection for another 2-4 weeks. If a volunteer tests positive or has symptoms of COVID-19, this period might be extended and they might be moved to additional clinical facilities as needed. Throughout the study period, participants would be tested extensively, with blood draws, stool samples and nasal swabs conducted on a daily or even more frequent basis. In addition to the risks of being infected with COVID-19, the experimental vaccine may have its own risks. We will not know in advance whether the experimental vaccine or controlled infection will enable volunteers to be protected against the virus in the future.

**Questionnaire**

1. **In what way did you first hear about the idea of a COVID-19 infection study?**

2. **What was your first reaction to the idea of a COVID-19 infection study?**

3. **When did you decide you wanted to enroll in a COVID-19 infection study?** *I.e. Is it a decision you made immediately or did you take some time to think about it?*

**Please indicate how important the following factors are in your decision to volunteer for a controlled COVID-19 infection study[1]:**

| | | | | | |
|---|---|---|---|---|---|
| **4. I want to help others and potentially save lives** | Strongly disagree | Somewhat disagree | Neither agree nor disagree | Somewhat agree | Strongly agree |
| **5. I feel helpless and this is a way to do something positive** | Strongly disagree | Somewhat disagree | Neither agree nor disagree | Somewhat agree | Strongly agree |

| | | | | | |
|---|---|---|---|---|---|
| **6. I was curious about COVID-19** | Strongly disagree | Somewhat disagree | Neither agree nor disagree | Somewhat agree | Strongly agree |
| **7. I wanted to be part of a clinical trial** | Strongly disagree | Somewhat disagree | Neither agree nor disagree | Somewhat agree | Strongly agree |
| **8. I wanted to contribute to the progress of medicine** | Strongly disagree | Somewhat disagree | Neither agree nor disagree | Somewhat agree | Strongly agree |
| **9. I wanted to find out more about my own health** | Strongly disagree | Somewhat disagree | Neither agree nor disagree | Somewhat agree | Strongly agree |
| **10. I wanted to receive financial reimbursement for participating** | Strongly disagree | Somewhat disagree | Neither agree nor disagree | Somewhat agree | Strongly agree |
| **11. I wanted to be guaranteed access to critical care should I be infected with COVID-19** | Strongly disagree | Somewhat disagree | Neither agree nor disagree | Somewhat agree | Strongly agree |
| **12. I am likely to be infected with COVID-19 anyway** | Strongly disagree | Somewhat disagree | Neither agree nor disagree | Somewhat agree | Strongly agree |

**13. Are there any other factors that influenced your decision to volunteer for a controlled COVID-19 infection study?**

**14. What are the three most important factors in your decision to participate?** *Please select three options*

❏ I want to help others and potentially save lives
❏ I feel helpless and this is a way to do something positive
❏ I was curious about COVID-19
❏ I wanted to be part of a clinical trial
❏ I wanted to contribute to the progress of medicine
❏ I wanted to find out more about my own health
❏ I want to receive the financial reimbursement for participating
❏ I wanted to be guaranteed access to critical care should I be infected with COVID-19
❏ I am likely to be infected with COVID-19 anyway

❏ Other factor not mentioned (please specify): ______________________________________

**15. Please indicate the extent to which you agree with the following statement: COVID-19 is of similar severity as influenza (the flu)**

| | | | | | | |
|---|---|---|---|---|---|---|
| 1<br>Strongly disagree | 2<br>Disagree | 3<br>Somewhat disagree | 4<br>Undecided | 5<br>Somewhat agree | 6<br>Agree | 7<br>Strongly agree |

**16. What do you think are the main risks and burdens of being in a controlled COVID-19 infection study?**

**a. Which of these risks and burdens concern you the most?**

**b. Which concern you the least?**

**17. What do you think are the main benefits of being in a controlled COVID-19 infection study?**

**18. How risky do you think a COVID-19 challenge study is overall, on a scale of 1 to 10?** This refers to the risk to your own health from participating — not a risk to others, and not including how likely you are to be infected with COVID-19 outside of a trial.

| | | | | | | | | | |
|---|---|---|---|---|---|---|---|---|---|
| 1<br>Not risky at all | 2 | 3 | 4 | 5<br>Moderately risky | 6 | 7 | 8 | 9 | 10<br>Extremely risky |

**19. What is your perception of the risk of developing conditions during a COVID-19 challenge trial that are severe enough to require critical care such as a ventilator?**

- 1 in 100,000 (0.001%)
- 1 in 50,000 (0.002%)
- 1 in 10,000 (0.01%)
- 1 in 5,000 (0.02%)
- 1 in 1,000 (0.1%)
- 1 in 100 (1%)
- 1 in 10 (10%)
- 1 in 5 (20%)

**20. How much care do you predict you would receive from the researchers if you were to become sick?**

| 1<br>None at all | 2<br>A little | 3<br>A moderate amount | 4<br>A lot | 5<br>A great deal |
|---|---|---|---|---|

**21. How much care do you think you have the right to receive from the researchers if you were to become sick?**

| 1<br>None at all | 2<br>A little | 3<br>A moderate amount | 4<br>A lot | 5<br>A great deal |
|---|---|---|---|---|

**22. How would you rate your risk of dying should you become infected with COVID-19 during the course of a challenge trial, if you were to participate?**

| 1<br>There is no risk to me | 2 | 3 | 4<br>There is a moderate risk to me | 5 | 6 | 7<br>The risk to me is extremely high |
|---|---|---|---|---|---|---|

**23. What is your perception of your risk of dying should you become infected with COVID-19 during a challenge trial, if you were to participate?**

- 1 in 100,000 (0.001%)
- 1 in 50,000 (0.002%)
- 1 in 10,000 (0.01%)
- 1 in 5,000 (0.02%)
- 1 in 1,000 (0.1%)
- 1 in 100 (1%)
- 1 in 10 (10%)
- 1 in 5 (20%)
- 1 in 3 (33.3%)

**24. Compared to your expected risk in participating in a COVID-19 challenge trial, to what extent would you be willing to accept greater risk as a participant if you believed the benefits to society would be significant?**

| 1<br>I would not accept any additional risk | 2 | 3 | 4<br>I would accept a moderately greater level of risk | 5 | 6 | 7<br>I would accept a seriously greater level of risk |
|---|---|---|---|---|---|---|

**25. What is the highest risk of death that you would accept to participate in a COVID-19 challenge trial if you believed such a trial was likely to confer significant benefits to society?**

- 1 in 100,000 (0.001%)
- 1 in 50,000 (0.002%)
- 1 in 10,000 (0.01%)
- 1 in 5,000 (0.02%)
- 1 in 1,000 (0.1%)
- 1 in 100 (1%)
- 1 in 10 (10%)
- 1 in 5 (20%)
- 1 in 3 (33.3%)

**26. How would you rate your risk of becoming infected with COVID-19 during the course of your everyday life in the next six months?**

| | | | | | | |
|---|---|---|---|---|---|---|
| 1<br>There is no risk to me | 2 | 3 | 4<br>There is a moderate risk to me | 5 | 6 | 7<br>The risk to me is extremely high |

**27. What is your perception of your risk of becoming infected with COVID-19 during the course of your everyday life in the next six months?**

- 1 in 100,000 (0.001%)
- 1 in 50,000 (0.002%)
- 1 in 10,000 (0.01%)
- 1 in 5,000 (0.02%)
- 1 in 1,000 (0.1%)
- 1 in 100 (1%)
- 1 in 10 (10%)
- 1 in 5 (20%)
- 1 in 3 (33%)
- 1 in 2 (50%)
- 3 in 4 (75%)
- 9 in 10 (90%)
- Almost certainly (95% or greater)

**28. How would you describe your risk of <u>not</u> being put on a ventilator or receiving other scarce treatments (such as dialysis machines) that you need to survive in your current living area because of the COVID-19 surge in demand for health services?**

| 1<br>There is no risk to me | 2 | 3 | 4<br>There is a moderate risk to me | 5 | 6 | 7<br>The risk to me is extremely high |
|---|---|---|---|---|---|---|

**29. What is your perception of your risk of <u>not</u> being put on a ventilator or receiving other scarce treatments (such as dialysis machines) that you need to survive in your current living area because of the COVID-19 surge in demand for health services?**

- 1 in 100,000 (0.001%)
- 1 in 50,000 (0.002%)
- 1 in 10,000 (0.01%)
- 1 in 5,000 (0.02%)
- 1 in 1,000 (0.1%)
- 1 in 100 (1%)
- 1 in 10 (10%)
- 1 in 5 (20%)
- 1 in 3 (33%)
- 1 in 2 (50%)
- 3 in 4 (75%)
- 9 in 10 (90%)
- Almost certainly (95% or greater)

**30. How would you rate your risk of dying from COVID-19 during the course of your everyday life in the next six months?**

| 1<br>There is no risk to me | 2 | 3 | 4<br>There is a moderate risk to me | 5 | 6 | 7<br>The risk to me is extremely high |
|---|---|---|---|---|---|---|

**31. What is your perception of your risk of dying from COVID-19 during the course of your everyday life in the next six months?**

- 1 in 100,000 (0.001%)
- 1 in 50,000 (0.002%)
- 1 in 10,000 (0.01%)
- 1 in 5,000 (0.02%)
- 1 in 1,000 (0.1%)
- 1 in 100 (1%)
- 1 in 10 (10%)
- 1 in 5 (20%)

**32. How many infections of COVID-19 do you think will have occurred worldwide, before 2021?**

- Between 2 million and 5 million infections
- Between 5 million and 20 million infections
- Between 20 million and 25 million infections
- Between 25 million and 50 million infections
- Between 50 million and 100 million infections
- Between 100 million and 250 million infections
- Between 250 million and 500 million infections
- Between 500 million and 750 million infections
- More than 750  million infections

**33. How many people do you think will die as a direct result of COVID-19 before 2021?**

- Under 20,000 deaths
- Between 20,000 and 50,000 deaths
- Between 50,000 and 100,000 deaths
- Between 100,000 and 250,000 deaths
- Between 250,000 and 500,000 deaths
- Between 500,000 and 750,000 deaths
- Between 750,000 and 1 million deaths
- Between 1 million and 2 million deaths
- Between 2 million and 5 million deaths
- Between 5 million and 10 million deaths
- More than 10 million deaths

**34. How many people do you think will die as a indirect result of COVID-19 before 2021?**
*This might include people who die of conditions that clinicians must neglect in order to prioritize COVID-19, halted economic development, ensuing famine, etc.*

- Under 20,000 deaths
- Between 20,000 and 50,000 deaths
- Between 50,000 and 100,000 deaths
- Between 100,000 and 250,000 deaths
- Between 250,000 and 500,000 deaths
- Between 500,000 and 750,000 deaths
- Between 750,000 and 1 million deaths
- Between 1 million and 2 million deaths
- Between 2 million and 5 million deaths
- Between 5 million and 10 million deaths
- More than 10 million deaths

**35. How likely do you think it is that your participation in a COVID-19 human challenge trial leads to accelerated vaccine development?**

| 1 | 2 | 3 | 4 | 5 | 6 | 7 |
|---|---|---|---|---|---|---|
| Extremely unlikely | Fairly unlikely | Somewhat unlikely | Neutral | Somewhat likely | Fairly likely | Extremely likely |

**36. How likely do you think it is that your participation in a COVID-19 human challenge trial leads to saving people's lives?**

| 1 | 2 | 3 | 4 | 5 | 6 | 7 |
|---|---|---|---|---|---|---|
| Extremely unlikely | Fairly unlikely | Somewhat unlikely | Neutral | Somewhat likely | Fairly likely | Extremely likely |

**37. Did you talk to anyone about whether you should join a potential human challenge study?**

- Yes
- No
- Prefer not to say

**a. If yes, who did you talk to?**
**b. What did they think about it?**

---

**Please indicate whether you believe the following statements are true or false.** This section does not require any additional outside research, and is simply designed to capture your current understanding of how a human challenge trial may work.

**38.** Participants in a human challenge trial would be exposed to the novel coronavirus (COVID-19):

- True
- False

**39.** Participants in a human challenge trial will be isolated from their household and general public in a specialized health facility

- True
- False

**40.** Participants in a human challenge trial might infect their family members with COVID-19

- True
- False

**41.** Participants are eligible for a human challenge trial if they have already tested positive for COVID-19

- True
- False

**42.** All participants will receive the candidate COVID-19 vaccine

- True
- False

**43. Have you ever participated in research studies before?**

| Never | Once | More than once | Often | Very Often |
|---|---|---|---|---|

**44. If you have participated in research studies before, tell us more about that experience.**

**a.** What was the purpose of the study?

**b.** Was your experience mostly positive or mostly negative?

**c.** How would this study be different from previous studies you have participated in?

**45. Have you ever donated blood before?**

| Never | Once | More than once | Often | Very Often |
|---|---|---|---|---|

**46. Have you ever donated a significant amount of money to charity?**

- Yes, I have occasionally
- Yes, I donate annually
- No, I have never donated to charity

**47. Are you a registered bone marrow donor?**

- Yes
- No
- Unsure/I don’t know
- Prefer not to say

**48. Are you a registered deceased organ donor?**

- Yes
- No
- Unsure/I don’t know
- Prefer not to say

**49. Have you ever been a living kidney or liver donor?**

- Yes, I am a kidney donor
- Yes, I am a liver donor
- No
- Prefer not to say

---

**50. If a COVID-19 human challenge trial moves forward and you are asked to participate, what is the likelihood you agree to participate?**

| | | | | | | |
|---|---|---|---|---|---|---|
| 1 | 2 | 3 | 4 | 5 | 6 | 7 |
| Extremely unlikely | Fairly unlikely | Somewhat unlikely | Neutral | Somewhat likely | Fairly likely | Extremely likely |

**51. If an effective treatment for COVID-19 is discovered, and a COVID-19 human challenge trial moves forward, what is the likelihood you agree to participate?**

| | | | | | | |
|---|---|---|---|---|---|---|
| 1 | 2 | 3 | 4 | 5 | 6 | 7 |
| Extremely unlikely | Fairly unlikely | Somewhat unlikely | Neutral | Somewhat likely | Fairly likely | Extremely likely |

**52. If a COVID-19 human challenge trial moves forward and you are asked to participate, what would make you NOT want to enroll in such a study?**

**53. How likely do you think it is that some people participating in the study might not tell the researchers things that could disqualify them from participating?** *For example, not fully revealing drug or alcohol consumption, or personal history that may exclude them from participating otherwise.*

| | | | | | | |
|---|---|---|---|---|---|---|
| 1 | 2 | 3 | 4 | 5 | 6 | 7 |
| Extremely unlikely | Fairly unlikely | Somewhat unlikely | Neutral | Somewhat likely | Fairly likely | Extremely likely |

**54. How do you think other people would perceive your enrollment in a COVID-19 challenge trial?**

| 1 | 2 | 3 | 4 | 5 | 6 | 7 |
|---|---|---|---|---|---|---|
| Very negatively | Fairly negatively | Somewhat negatively | Neutral | Somewhat positively | Fairly positively | Very positively |

**55. Where others (e.g., their parents or colleagues) may be upset about your enrollment, what would be helpful, if anything, for us or the trial team to do to pre-empt or address these reactions?** *For example, providing you with information about the study to share with others?*

**56. Please indicate whether any of your personal connections have been infected with COVID-19.**
*A severe case is defined here as requiring hospitalization or admission to an intensive care unit (ICU).*

| Personal Connection | COVID-19 Status |
|---|---|
| Partner/Spouse | N/A Mild Severe Fatal |
| Parent | N/A Mild Severe Fatal |
| Child | N/A Mild Severe Fatal |
| Sibling | N/A Mild Severe Fatal |
| Close friend | N/A Mild Severe Fatal |
| Work colleague | N/A Mild Severe Fatal |
| Housemate | N/A Mild Severe Fatal |
| Remote friend | N/A Mild Severe Fatal |
| Extended family | N/A Mild Severe Fatal |

| | |
|---|---|
| Classmate | N/A Mild Severe Fatal |
| Neighbor | N/A Mild Severe Fatal |
| Other: | N/A Mild Severe Fatal |

**57. Are any of your personal connections considered at-risk of becoming infected with COVID-19 or high-risk?** *At-risk might include healthcare workers, other front-line workers or people residing in areas heavily burdened by COVID-19.*
*High-risk might include people who are 70 or older, are pregnant, or have an underlying condition that increases their risk of COVID-19 complications.*

- Yes
- No
- Unsure/I don't know
- Prefer not to say

**a.** If yes, please specify below. Complete for all that apply:

| Personal Connection | COVID-19 Risk Designation | | Lives in Household | |
|---|---|---|---|---|
| Partner/Spouse | At-risk | High- risk | Yes | No |
| Parent | At-risk | High- risk | Yes | No |
| Child | At-risk | High- risk | Yes | No |
| Sibling | At-risk | High- risk | Yes | No |
| Close friend | At-risk | High- risk | Yes | No |
| Work colleague | At-risk | High- risk | Yes | No |
| Housemate | At-risk | High- risk | Yes | No |
| Remote friend | At-risk | High- risk | Yes | No |
| Extended family | At-risk | High- risk | Yes | No |
| Classmate | At-risk | High- risk | Yes | No |
| Neighbor | At-risk | High- risk | Yes | No |

| | | |
|---|---|---|
| Other: | At-risk High- risk | Yes No |

**Please indicate the extent to which you agree with the following statements[2]:**

**58.** I have confidence in doctors and medical professionals

| | | | | | | |
|---|---|---|---|---|---|---|
| 1 Strongly disagree | 2 Disagree | 3 Somewhat disagree | 4 Undecided | 5 Somewhat agree | 6 Agree | 7 Strongly agree |

**59.** I trust doctors' judgement about my medical care

| | | | | | | |
|---|---|---|---|---|---|---|
| 1 Strongly disagree | 2 Disagree | 3 Somewhat disagree | 4 Undecided | 5 Somewhat agree | 6 Agree | 7 Strongly agree |

**60.** I have confidence in scientific researchers

| | | | | | | |
|---|---|---|---|---|---|---|
| 1 Strongly disagree | 2 Disagree | 3 Somewhat disagree | 4 Undecided | 5 Somewhat agree | 6 Agree | 7 Strongly agree |

***B. Demographic Information***

| | |
|---|---|
| **61. City** | |
| **62. State** | |
| **63. Post (ZIP) Code** | |
| **64. Country** | |
| **65. Year of Birth** | |
| **66. Height** | cm OR ft inches |
| **67. Weight** | kg OR pounds |

| | |
|---|---|
| **68. Gender** | Male<br>Female<br>Prefer to self describe as ____________ (non-binary, gender-fluid, agender, please specify)<br>Prefer not to say |
| **69. What is your marital status?** | Single (never married)<br>Married, or in a domestic partnership<br>Widowed<br>Divorced<br>Separated |

**70. Please specify your ethnicity (select all that apply):**

- African
- Asian (East Asian)
- Asian (South Asian)
- Caucasian (Non-Hispanic)
- Carribean
- Latino or Hispanic
- Native American
- Native Hawaiian or Pacific Islander
- Other/Unknown
- Prefer not to say

**71. What is the highest degree or level of school you have completed?**
*If currently enrolled, the highest degree received.*

- Less than high school equivalent
- High school graduate, diploma or the equivalent (for example: GED)
- Some college credit, no degree
- Trade/technical/vocational training
- Associate degree
- Bachelor’s degree
- Master’s degree
- Professional degree
- Doctoral degree

**72. What is your work status? (select all that apply)**

- Employed
- Self-employed/Freelance
- Interning
- Part-time

- Unemployed- Looking for work
- Unemployed – Not looking for work
- Homemaker
- Studying
- Military/Armed Forces
- Retired
- Not able to work
- Other

**73. Are you currently working from home?**

- Yes, full-time
- Yes, part-time (and part-time from my workplace)
- No

**74. Are you classified as an essential worker by your government?** *(Essential sectors could include health and social care, education and childcare, key public services—justice system, mortuaries, journalists —local and national government, those involves in food or other necessary goods production, distribution or sale and delivery, public safety and national security, transport, or utilities, communication and financial services)*

- Yes
- No
- I'm not sure

**75. What is your annual household income?**

- Less than $25,000 USD
- $25,000 - $50,000 USD
- $50,000 - $100,000 USD
- $100,000 - $200,000 USD
- More than $200,000 USD
- Prefer not to say

**76. Do you currently have health insurance?**

- Yes
- No
- I'm not sure

  **a. If yes, who pays for your health insurance? (Check all that apply)**

- Government
- Current employer
- Former employer
- Self-funded
- Other: ________

**77. Do you have children? If yes, how many?**

- Yes, I have ___ children
- No

**78. Do you have dependents? If yes, how many?** *Dependents refer to people you financially support and would not include a spouse that generates their own income but may include children or elderly parents, etc.*

- Yes, I have ___ dependents (include the children from your answer above if they are dependent on you)
- No

**79. How many other people do you share your household with?**

| | | | | | | | | |
|---|---|---|---|---|---|---|---|---|
| 0 | 1 | 2 | 3 | 4 | 5 | 6 | 7 | 8+ |

**80. How many rooms are there in your household?** *This includes kitchens, living rooms, bedrooms, bathrooms, etc.*

| | | | | | | | | |
|---|---|---|---|---|---|---|---|---|
| 0 | 1 | 2 | 3 | 4 | 5 | 6 | 7 | 8+ |

**81. Do you have any of the following medical issues? (Select all that apply)**

- Asthma
- Blood disorders (such as sickle cell disease or taking blood thinners (*anticoagulants*))
- Cancer
- Chronic kidney disease
- Chronic liver disease
- Compromised immune system
- Currently pregnant or a recent pregnancy in the last two weeks
- Endocrine disorders, including diabetes
- Heart disease
- High blood pressure
- Lung disease, including asthma
- Metabolic disorders
- Neurological and neurologic and neurodevelopment conditions

**82. Do you currently smoke tobacco?**

- Yes, daily
- Yes, but not daily
- No, but I used to smoke daily
- No, but I used to smoke less than daily
- No, I have never smoked tobacco
- I'd rather not say

**83. Do you currently smoke marijuana?**

- Yes, daily
- Yes, but not daily
- No, but I used to smoke daily
- No, but I used to smoke less than daily
- No, I have never smoked marijuana
- I'd rather not say

**84. Do you currently use vaping products?**

- Yes, daily
- Yes, but not daily
- No, but I used to use them daily
- No, but I used to use them less than daily
- No, I have never used vaping products
- I'd rather not say

### Frameworks/Tools:

**The HEXACO Personality Index (10-15 minutes)**

- A measure of the six major dimensions of personality
- https://hexaco.org/

**DOSPERT Scale (15-20 minutes)**

- Assesses both conventional risk attitudes (defined as the reported level of risk taking) and perceived-risk attitudes (defined as the willingness to engage in a risky activity as a function of its perceived riskiness) in five commonly encountered content domains, i.e., ethical, financial (further decomposed into gambling and investment), health/safety, social, and recreational decisions.
- Weber, E. U., Blais, A.-R., & Betz, N. (2002). A domain-specific risk-attitude scale: Measuring risk perceptions and risk behaviors. Journal of Behavioral Decision Making, 15, 263-290.
- Blais, A.-R., & Weber, E. U. (2006). A domain-specific risk-taking (DOSPERT) scale for adult populations. *Judgment and Decision Making*, *1*(1).

---

[1] Oguti, B., Gibani, M., Darlow, C., Waddington, C. S., Jin, C., Plested, E., Campbell, D., Jones, C., Darton, T. C., & Pollard, A. J. (2019). Factors influencing participation in controlled human infection models: a pooled analysis from six enteric fever studies. Wellcome Open Research, 4, 153.

[2] Partially adapted from Kelly, J. J., Njuki, F., Lane, P. L., & McKinley, R. K. (2005). Design of a Questionnaire to Measure Trust in an Emergency Department. *Academic Emergency Medicine*, *12*(2), 147–151.
